# Clinical Performance of Cas13a-based Point-of-Care Lateral Flow Assay for Detecting *Neisseria gonorrhoeae*

**DOI:** 10.1101/2024.03.01.24303603

**Authors:** Lao-Tzu Allan-Blitz, Gabriela Sanders, Palak Shah, Gordon Adams, Jana Jarolimova, Kevin Ard, John A. Branda, Jeffrey D. Klausner, Pardis C. Sabeti, Jacob E. Lemieux

**Affiliations:** Division of Global Health Equity: Department of Medicine, Brigham and Women’s Hospital, Boston, MA; Broad Institute of Massachusetts Institute of Technology and Harvard, Boston, MA; Division of Infectious Diseases: Department of Medicine, Massachusetts General Hospital, Boston, MA; Department of Pathology, Massachusetts General Hospital, Boston, MA; Department of Population and Public Health Sciences, Keck School of Medicine, University of Southern California, Los Angeles, CA

## Abstract

**Background:** Diagnosis of *Neisseria (N.) gonorrhoeae* is dependent on nucleic acid amplification testing (NAAT), which is not available in resource-limited settings where the prevalence of infection is highest. Recent advances in molecular diagnostics leveraging the high specificity of CRISPR enzymes can permit field-deployable, point-of-care lateral flow assays. We previously reported on the development and *in vitro* performance of a lateral flow assay for detecting *N. gonorrhoeae*. Here we aimed to pair that assay with point-of-care DNA extraction techniques and assess the performance on clinical urine specimens.

**Methods:** We collected an additional urine specimen among individuals enrolling in an ongoing clinical trial at the Massachusetts General Hospital Sexual Health Clinic who presented with symptoms of urethritis or cervicitis (urethral or vaginal discharge, dysuria, or dyspareunia). We then assessed thermal, detergent, and combination DNA extraction conditions, varying the duration of heat at 95°C and concentration of Triton X. We assessed the efficacy of the various DNA extraction methods by quantitative polymerase chain reaction (qPCR). Once an extraction method was selected, we incubated samples for 90 minutes to permit isothermal recombinase polymerase amplification. We then assessed the performance of lateral flow Cas13a-based detection using our previously designed *por*A probe and primer system for *N. gonorrhoeae* detection, comparing lateral flow results with NAAT results from clinical care.

**Results:** We assessed DNA extraction conditions on 3 clinical urine specimens. There was no consistent significant difference in copies per microliter of DNA obtained using more or less heat. On average, we noted that 0.02% triton combined with 5 minutes of heating to 95°C resulted in the highest DNA yield, however, 0.02% triton alone resulted in a quantity of DNA that was above the previously determined analytic sensitivity of the assay. Given that detergent-based extraction is more easily deployable, we selected that as our method for extraction. We treated 23 clinical specimens with 0.02% triton, which we added to the Cas13a detection system. We ran all lateral flow detections in duplicate. The Cas13a-based assay detected 8 of 8 (100%) positive specimens, and 0 of 15 negative specimens.

**Conclusion:** Using point-of-care DNA extraction, isothermal amplification, and Cas13a-based detection, our point-of-care lateral flow *N. gonorrhoeae* assay correctly identified 23 clinical urine specimens as either positive or negative. Further evaluation of this assay among larger samples and more diverse sample types is warranted.

## Introduction

Despite a high and increasing prevalence of infection worldwide,^1^ with the highest burden of disease in resource-limited settings,^2,3,4^ the diagnosis of *Neisseria (N.) gonorrhoeae* is predominantly dependent on nucleic acid amplification testing (NAAT).^5^ ^6^ NAAT historically has required both laboratory infrastructure and higher costs, which preclude its use in low-resource areas. *N. gonorrhoeae* culture, while low-cost, is too time and labor intensive to be clinically useful,^7^ and also requires laboratory infrastructure unavailable in many areas where the disease is most prevalent.

Thus, low-resource settings utilize syndromic management, in which patients with symptoms of urethritis and cervicitis are treated empirically. Syndromic management, however, misses a high proportion of asymptomatic cases,^8–11^ and contributes to the emergence of antimicrobial resistance in *N. gonorrhoeae* via the overuse of antibiotics.^12,13^ Consequently, low-resource areas may have some of the highest rates of antimicrobial-resistant *N. gonorrhoeae* infection ^14–16^ - an urgent global public health threat.^17^

Novel point-of-care diagnostics for *N. gonorrhoeae* are increasingly available.^18,19^ ^20^ Further, some of those tests appear to be both feasible and acceptable in low-resource settings.^21^ However, few have met the World Health Organization (WHO) standards for point-of-care tests: real-time connectivity, ease of specimen collection, affordable, sensitive, specific, user-friendly, rapid and robust, equipment free/environmentally friendly, deliverable to end-users.^22^ We recently reported on the development of a Cas13a-based point-of-care test for both detecting *N. gonorrhoeae* and predicting antimicrobial resistance.^22,23^ That assay demonstrated promising *in vitro* performance on purified isolates, but used DNA extraction techniques which required laboratory training and infrastructure, and has not been evaluated on clinical specimens. We now aimed to evaluate the performance of that assay on clinical urine specimens using point-of-care DNA extraction.

## Methods

### Ethics statement

The Massachusetts General Brigham Institutional Review Board gave ethical approval for this study, as well as the secondary use of clinical specimens.

### Specimen Collection

We enrolled participants presenting with symptoms of urethritis or cervicitis to the Massachusetts General Hospital Sexual Health Clinic as a part of an ongoing clinical trial (NCT05564299). We enrolled patients between March and September 2023 if they reported urethral or virginal discharge, dysuria, or dyspareunia. Additional inclusion criteria included: 18 years or older, not known to be pregnant at the time of enrollment, had no known exposure to *N. gonorrhoeae* or *Chlamydia trachomatis* within the past 6 weeks, had no concurrent symptoms at extragenital sites, and were willing to provide informed consent.

Among eligible patients who provided informed consent, we collected one additional urine specimen from male patients for testing with our Cas13-based assay. All participants received standard-of-care diagnostics and therapeutics, which included gram stain, NAAT, and culture with susceptibility testing using standard methods. We considered specimens to be *N. gonorrhoeae* positive if standard clinical diagnostics were positive. Neither providers nor patients were informed of the results of Cas13a-based testing.

We aliquoted clinical urine specimens using a sterile pipette into CryoTube Vials, each with 1.5 mL of urine specimen, and assigned a unique study identifier to each specimen. We stored all specimens at −80° C within 30 minutes of aliquoting.

### DNA Extraction

DNA extraction was performed by several methods. Thermal-based nucleic acid extraction has been recently used for SARS-CoV-2 qPCR assays,^24,25^ while prior work demonstrated Triton X can disrupt cell membranes while simultaneously solubilizing membrane proteins.^26^ We thus evaluated both thermal-extraction and extraction using Triton X with or without heat. In the heat alone conditions, we incubated urine samples at 95C for 30s, 1 minute, 2 minutes, 5 minutes, 10 minutes, or 15 minutes either directly from the working stock or after either centrifugation for 5 minutes at 8000 rpm. In the Triton X conditions, we centrifuged urine samples for 5 minutes at 8000 rpm, and the resulting pellet was resuspended in 0.01%, 0.02%, or 0.1% Triton X. We also added Triton X directly to unspun urine samples at a final concentration of either 0.01%, 0.02%, or 0.1%. In addition, we treated urine samples with both heat and the varying concentrations of Triton X.

We assessed the efficacy of the various DNA extraction methods by quantitative polymerase chain reaction (qPCR). The forward and reverse primer sequences for the *Neisseria gonorrhoeae gyr*A gene were 5’ GCGACGGCCTAAAGCCAGTG 3’ and 5’ GTCTGCCAGCATTTCATGTGAG 3’, respectively. qPCR reaction mixtures contained 1x FastStart SYBR Green Master Mix (Sigma Aldrich, United States), 0.5 µM of each primer, and DNA template in a 1:9 template to master mix ratio. We adjusted the final qPCR reaction volume to 10 µL with nuclease-free water, and loaded in triplicate on a 384-well plate, which we ran on a QuantStudio 6 (Applied Biosystems, United States) under the following cycle conditions: heat activation at 95° C for 10 minutes, 40 cycles of a denaturing step at 95° C for 15 seconds, an annealing step at 60° C for 1 minute, and an extension step at 72° C, followed by a final extension step at 68° C for 2 minutes. We collected amplification data during the second extension stage and analyzed these data using the Standard Curve module of the Applied Biosystems Analysis Software. We then quantified DNA against a standard curve.

### Lateral flow SHERLOCK assay

We performed SHERLOCK reactions using 45 nM C2c2 *Lwa*Cas13a (GenScript Biotech Corp, United States) resuspended in 1x storage buffer (SB: 50 mM Tris (pH 7.5), 600 mM KCl, 5% glycerol, and 2mM dithiothreitol) such that the resuspended protein was at 2.25 μM, 1 U/μL murine RNase Inhibitor (New England Biosciences, United States), 10 U/μL T7 RNA polymerase (Lucigen, United States), 1 μM of a biotinylated FAM reporter (Integrated DNA Technologies, United States), 1x SHINE Buffer (SHINE: 20 mM HEPES (4-(2-hydroxyethyl)-1-piperazineethanesulfonic acid) (pH 8.0), 60 mM KCl, and 5% polyethylene glycol), and 2 mM of each rNTP (New England Biosciences, United States). We then added crRNA, final concentration 22.5 nM, and primers, final concentration 320 nM.

We rehydrated the TwistAmp Basic Kit lyophilized recombinase polymerase amplification (RPA) pellets (1 pellet per 73.42 μL master mix volume) using the prepared master mix. We then added 14 mM MgAOc (TwistDx, United Kingdom) after resuspension to activate the RPA pellets. Subsequently, we subdivided the master mix for each guide-primer set pair being analyzed, to which we added 22.5 nM gRNA (Integrated DNA Technologies, United States) and 320 nM each of the RPA primers (Integrated DNA Technologies, United States).

We then incubated the samples at 37 °C for 90 minutes, following which we added 80 μL of HybriDetect assay buffer (Milennia Biotec, Germany) to each sample in a 1:4 dilution along with a HybriDetect lateral flow strip (Milennia Biotec, Germany). We took images using a smartphone camera 3-5 minutes after the strips were added.

## Results

### DNA Extraction

In total, we assessed DNA extraction conditions on 3 clinical urine specimens. We used separate aliquots for DNA extraction experiments and *N. gonorrhoeae* detection experiments. Combination thermal and detergent-based conditions resulted in successful DNA extraction (Figure 1). There was no consistent significant difference in copies per microliter of DNA obtained using more or less heat. On average, we noted that 0.02% triton combined with 5 minutes of heating to 95°C resulted in the highest DNA yield.

**Figure 1:**
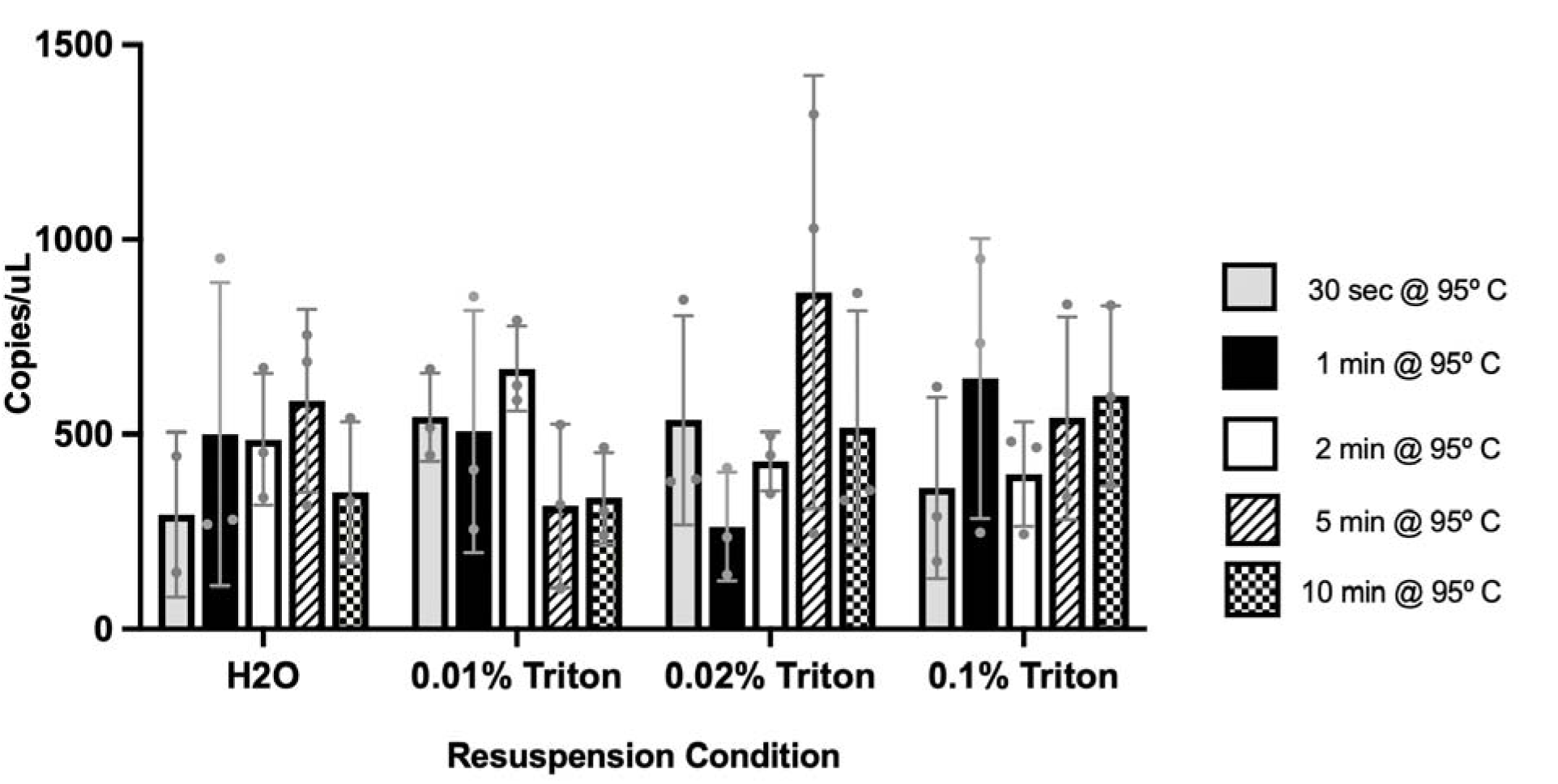
Quantification of *N. gonorrhoeae* DNA Extraction Using Combinations o Thermal and Chemical Lysis. The figure shows copies per microliter of *N. gonorrhoeae* DNA that resulted from combination thermal and detergent extraction conditions. Dots represent distribution of data run in triplicate.

We then assessed the viability of detergent-based extraction using pelleted DNA after centrifugation, which showed promising levels of DNA extraction. Subsequently, we compared single detergent extraction without centrifugation with combined detergent-heat extraction (Figure 2). Again, heating to 95°C for 5 minutes resulted in the highest yield of DNA regardless of detergent added. However, we noted impaired fluorescent signals with longer thermal durations. Further, 0.02% triton consistently resulted in DNA quantities above the previously determined analytic sensitivity of the assay.^23^ Additionally, single detergent extraction permits easier field deployment. Thus, we elected to use a single detergent-based extraction with 0.02% Triton. Of note, we observed up to 1,000 DNA copies per microliter of *N. gonorrhoeae* DNA even without the addition of a detergent.

**Figure 2:**
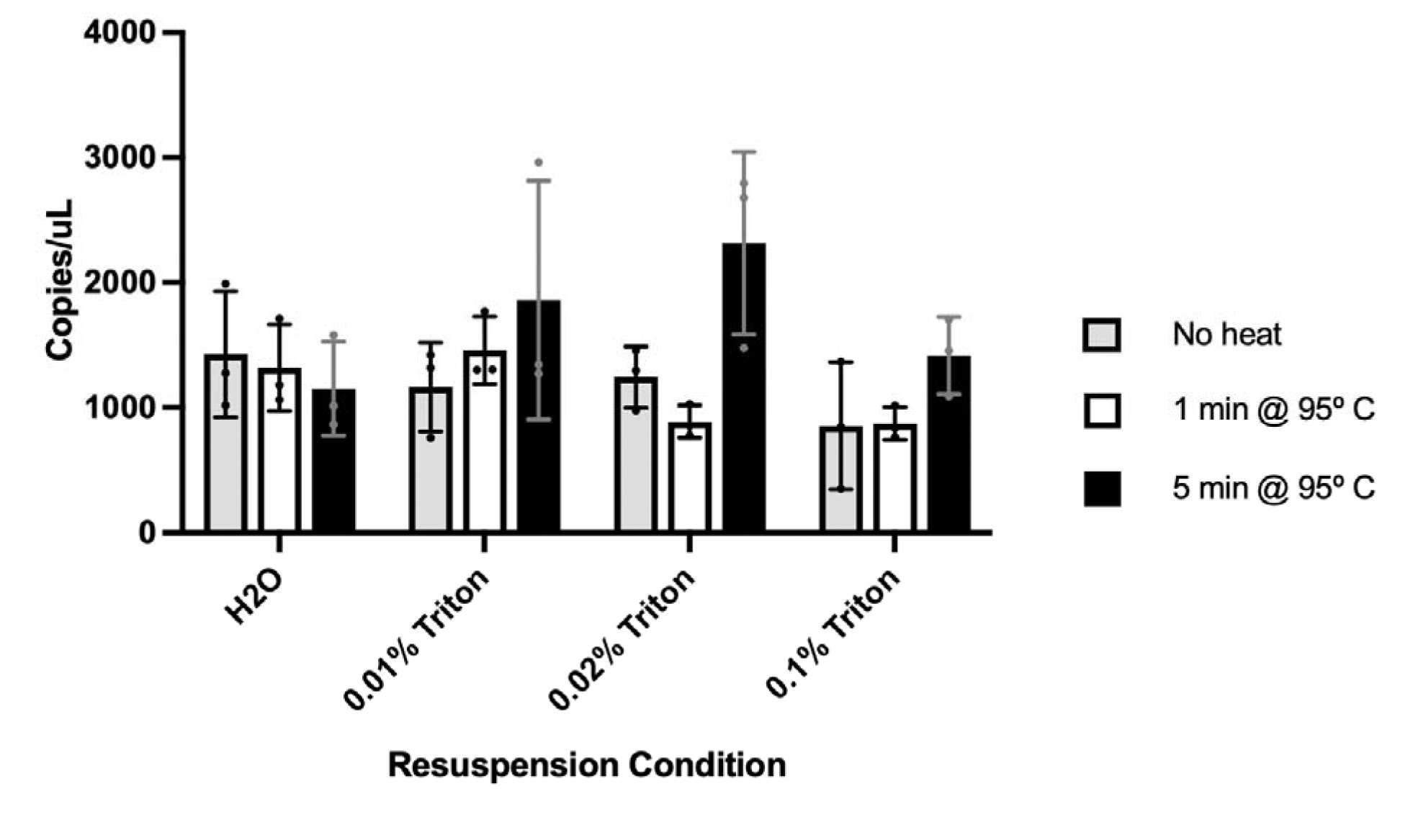
Quantification of *N. gonorrhoeae* DNA Extraction Comparing Detergent Only Extraction with Combined Detergent and Thermal Extraction. The figure shows the copies per microliter of *N. gonorrhoeae* DNA extracted using either detergent alone or detergent plus heat (95°C for 1 minute or 5 minutes).

### Lateral Flow N. gonorrhoeae Detection on Clinical Specimens

We assessed the performance of the lateral flow assay on 23 clinical urine specimens. We stored aliquots of clinical urine specimens at −80°C. For each detection reaction, we used a fresh aliquot, to which we added 0.02% triton X for point-of-care DNA extraction. We used that mixture as the target for lateral flow detection. We ran lateral flow detections in duplicate. The Cas13a-based assay detected 8 of 8 (100%) positive specimens, and 0 of 15 negative specimens (Figure 3).

**Figure 3:**
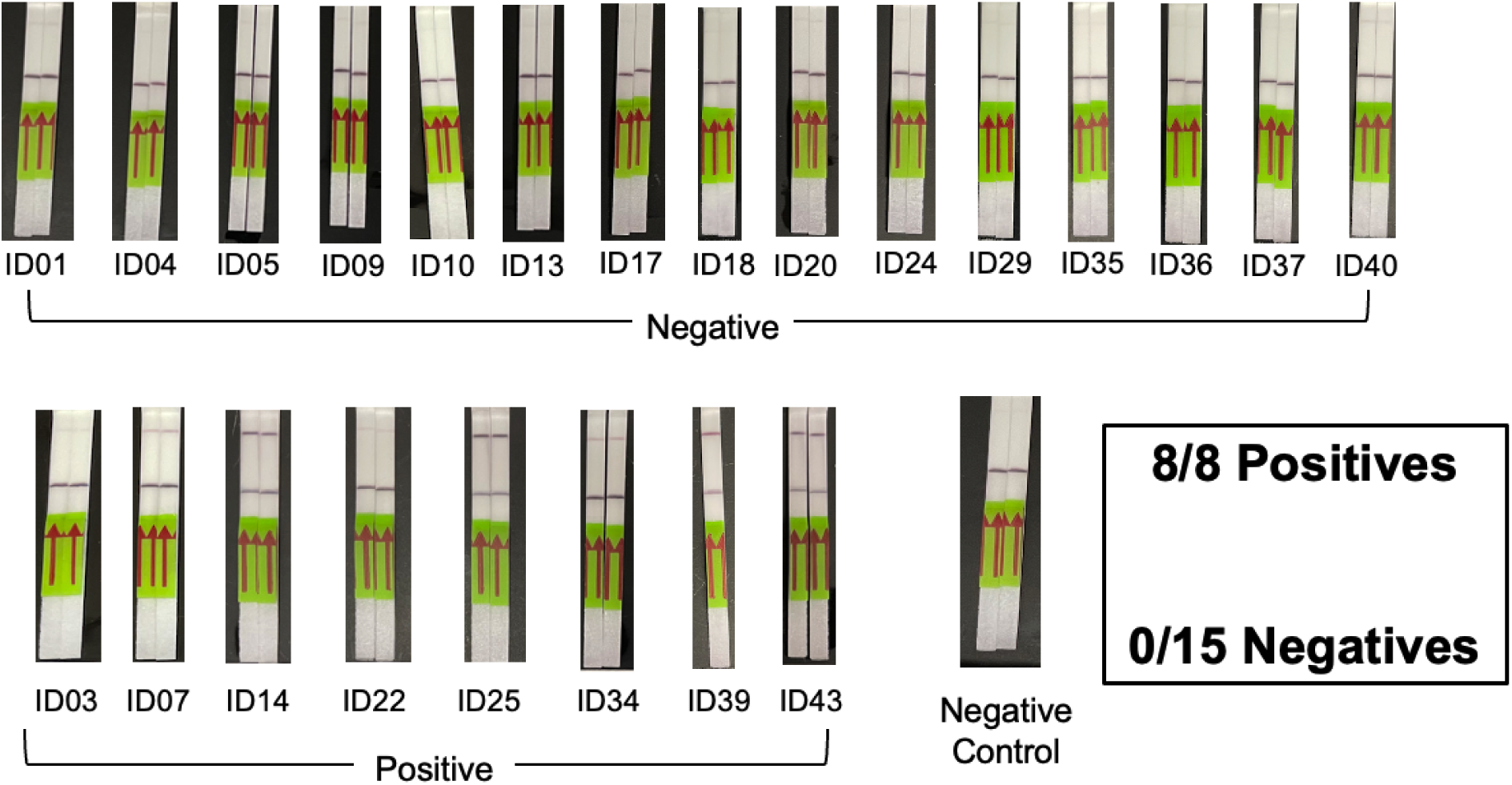
Performance of Cas13a-based lateral flow detection of *N. gonorrhoeae* on clinical urine specimens using detergent-based DNA extraction. The figure shows the results of our point-of-care *N. gonorrhoeae* lateral flow detection using detergent-based DNA extraction and incubation for 90 minutes. All 23 specimens were correctly classified as either positive or negative by the lateral flow platform.

As a follow-up experiment, we randomly selected two positive specimens and repeated the detection reaction varying the incubation duration: 10 minutes, 30 minutes, 60 minutes, and 90 minutes. We observed faint detection after 10 minutes and 30 minutes. For both specimens, we noted detection clearly visible after 60 minutes, but not as pronounced as after 90 minutes (Figure 4).

**Figure 4:**
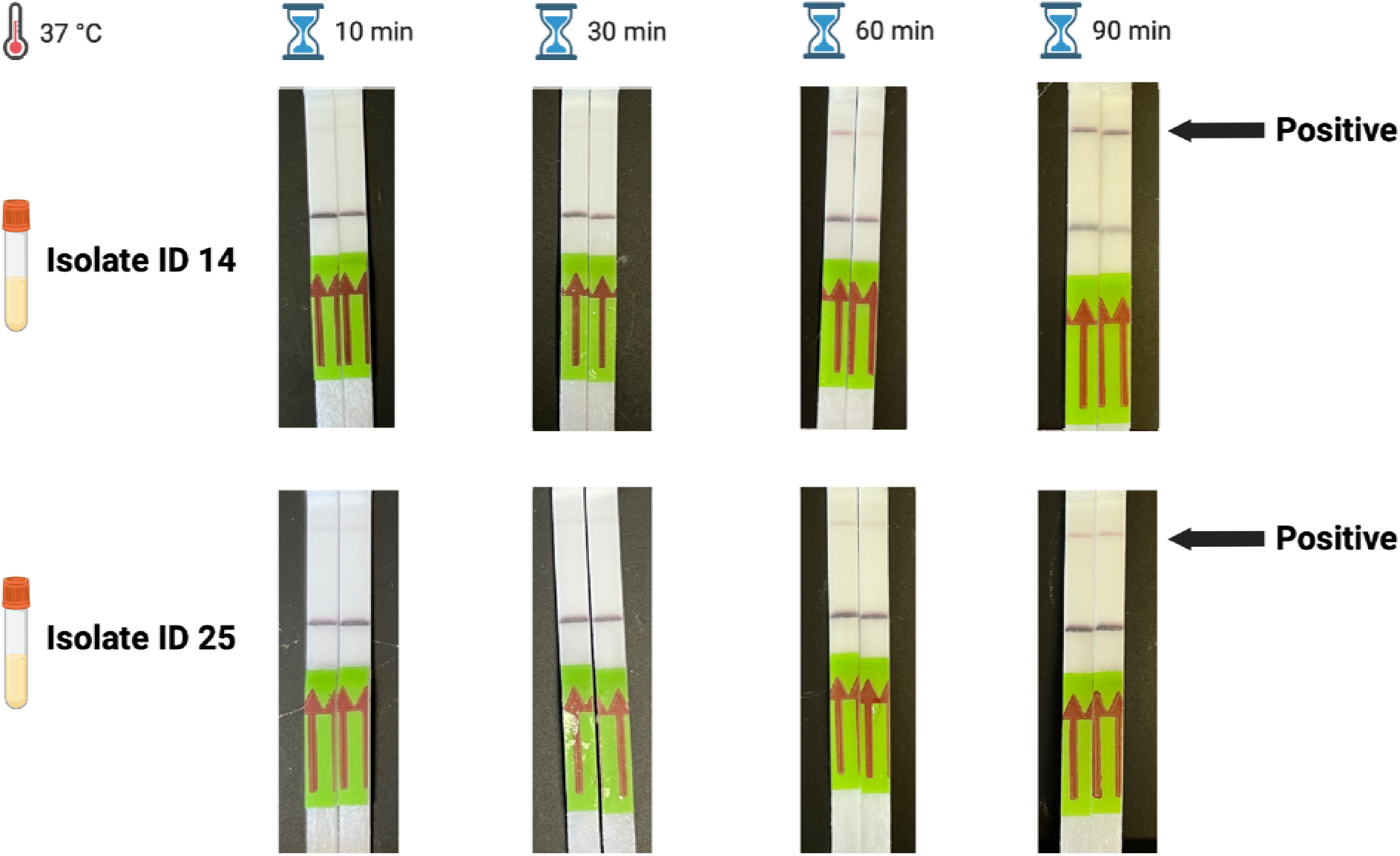
Varying Incubation Lengths for Isothermal Recombinant Polymerase Amplification and Cas13a-based Lateral Flow Point-of-Care Detection of Two *N. Gonorrhoeae* Specimens. The figure shows the results of 10, 30, 60, and 90 minute incubations for isothermal recombinant polymerase amplification permitting Cas13a-based detection of two *N. gonorrhoeae* clinical specimens using point-of-care DNA extraction and a lateral flow platform.

## Discussion

We report on the performance of a Cas13a-based point-of-care lateral flow *N. gonorrhoeae* detection assay on clinical urine specimens. Our assay showed 100% agreement with standard-of-care NAAT results. Cas13a detection paired with POC DNA extraction has the potential to transform the care of sexually transmitted infections in resource-limited settings where insufficient laboratory infrastructure exists to support NAAT.

In order to facilitate point-of-care detection, we evaluated rapid DNA extraction using different detergents with or without heat. Detergent-based extraction, without heat, may be more easily deployed in low-resource settings where access to a heat source is limited. Our results indicate that extraction using Triton X alone, while not as efficient as column-based solid phase extraction, was sufficient for pathogen detection when combined with RPA and Cas13a. When used with detergent-based extraction, our assay was sensitive enough to detect specimens with DNA concentrations of 14 copies per microliter with a 90-minute incubation period.

An important consideration for this lateral flow detection system is that the concentration of DNA within a sample appears to directly impact the intensity of the test band. Two specimens tested (ID03 and ID22) had faint positive bands, which, while recognized as positive in a laboratory setting, may be incorrectly classified in real-world contexts.

Further work should explore optimization of the platform using different concentrations of reporter and sample in order to maximize band intensity. Machine-learning tools for image segmentation and pixel quantification may also support interpretation of lateral flow results, supporting clinical use of such assays.^27–29^

We also evaluated the performance of our system using shorter incubation times. While there may be some reduction in incubation time possible as evidenced by the detection in two specimens at 60 minutes, we suspect that optimal performance of this platform may require longer incubation times. To meet the standards of the WHO REASSURED criteria,^22^ more rapid amplification will be required. Additional considerations for increasing the speed of amplification include other isothermal methods, such as loop mediated amplification,^30^ helicase-dependent amplification,^31^ and strand-displacement amplification.^32^ Optimized DNA extraction methods may facilitate more rapid amplification by making more DNA available.

## Limitations and Future Directions

Our results have several limitations. First, we tested our assay on a small number of urine specimens. Additionally, the urine samples were frozen prior to analysis, which might affect assay performance. Further, based on the criteria through which participants were enrolled, all urine samples processed were from men. Given the promising performance of the assay, evaluation among larger and more diverse samples is warranted. This was a single pathogen detection system. Simultaneous detection of multiple pathogens (e.g., *Chlamydia trachomatis*) will be needed to supplant syndromic management. Additionally, incorporation of molecular targets predicting antimicrobial resistance has the potential to facilitate resistance-guided therapy in low-resource settings. Given the consequences of untreated and inadequately treated *N. gonorrhoeae* infection, which include pelvic inflammatory disease,^33^ infertility,^34^ neonatal blindness,^35^ and an increased risk for the acquisition of HIV infection,^36,37,38^ we feel this platform is a promising step towards improving care of *N. gonorrhoeae* infection globally.

## Conclusion

Using point-of-care DNA extraction, isothermal amplification, and Cas13a-based detection, our point-of-care lateral flow *N. gonorrhoeae* assay correctly identified 23 clinical urine specimens as either positive or negative. Further evaluation of this assay among larger samples and more diverse sample types is warranted.

## Disclosures

P.C.S. is a co-founder of, shareholder in, and consultant to Sherlock Biosciences and Delve Bio, as well as a board member of and shareholder in Danaher Corporation. J.E.L previously served as a consultant to SHERLOCK Biosciences. J.B. has received research funding from Analog Devices Inc., Zeus Scientific, Immunetics, Pfizer, DiaSorin and bioMerieux, and has been a paid consultant to Flightpath Biosciences, Tarsus Pharmaceuticals, T2 Biosystems, DiaSorin, and Roche Diagnostics. J.J. has received in-kind research support from binx health. The remaining authors have nothing to disclose.

## Data Availability

All data produced in the present study are available upon reasonable request to the authors

## Acknowledgements and Funding

This work was supported in part by the Massachusetts General Hospital Department of Medicine Innovation Program grant to R.H.G, NIH NIAID U19AI110818 to P.C.S, grants 2019123 and 2021287 from the Doris Duke Charitable Foundation to J.E.L., and a small project award from the American S see exually Transmitted Disease Association to J.J.

## Notes

### Author Declarations

The Massachusetts General Brigham Institutional Review Board gave ethical approval for this work, as well as the secondary use of clinical specimens.

